# Measurement of Bone Marrow Tumour Burden and Minimal Residual Disease in Waldenstrom’s Macroglobulinemia through Cell-free Whole Genome Sequencing

**DOI:** 10.1101/2025.05.11.25326925

**Authors:** Signy Chow, Dor D. Abelman, Arnavaz Danesh, Stephanie Pedersen, Ellen Nong Wei, David S. Scott, Adam Suleman, Kim Roos, Christine I. Chen, Neil Berinstein, Suzanne Trudel, Trevor J. Pugh

**Affiliations:** Sunnybrook Research Institute, University of Toronto, Toronto, ON, Canada; Odette Cancer Centre, Sunnybrook Health Sciences Centre, Toronto, ON, Canada; Division of Medical Oncology and Hematology, Temerty Faculty of Medicine, University of Toronto, Toronto, ON, Canada; Department of Medical Biophysics, University of Toronto, Toronto, ON, Canada; Princess Margaret Cancer Centre, University Health Network, Toronto, ON, Canada; Ontario Institute for Cancer Research, Toronto, ON, Canada

## Abstract

**Purpose:** We assessed the utility of blood cell-free DNA (cfDNA) whole genome sequencing (cfWGS) for minimal residual disease (MRD) monitoring in Waldenstrom’s macroglobulinemia (WM) by comparing this to 1) targeted panel sequencing of 27 genes of interest in WM and targeted capture of immunoglobulin gene rearrangements in blood and bone marrow 2) Multiplex-PCR of immunoglobulin loci followed by Illumina sequencing (clonoSEQ).

**Experimental design:** Samples were collected from 7 patients on a clinical trial who were treated uniformly with chemoimmunotherapy and Bruton’s Tyrosine Kinase inhibitor (BTKi). Samples were collected prior to starting treatment and at clinical timepoints up to 18 months. MRD detection technologies were compared across all timepoints.

**Results:** cfWGS was superior to both in-house targeted panel sequencing on cfDNA and clinical NGS in peripheral blood (PB) cells, using clinical bone marrow (BM) NGS as a standard. Tumor burden measured by cfWGS reflected MRD counts by clonoSEQ in BM.

**Conclusions:** cfWGS may be a valuable non-invasive alternative to bone marrow testing in WM patients who require close follow up and provides greater sensitivity than targeted panel sequencing of cfDNA.

**Statement of Translational Relevance:** Whole-genome sequencing in cell-free DNA (cfWGS) is a highly sensitive marker of minimal residual disease that has application as a biomarker in clinical trials. cfWGS more accurately reflects bone marrow tumor burden than other available non-invasive measures to date. Further exploration is warranted to determine its full potential for use in cancer diagnostics and research.

## Introduction

Waldenstrom’s macroglobulinemia (WM) is a rare indolent lymphoma. Diagnosis requires a characteristic infiltrate of lymphoplasmacytic lymphoma (LPL) cells in the bone marrow (BM) and detection of a monoclonal IgM paraprotein (1). First-line treatment for WM consists of either chemoimmunotherapy or a targeted therapy with a Bruton Tyrosine Kinase inhibitor (BTKi). Choice of therapy depends on patient and disease factors, including specific somatic alterations in *MYD88, CXCR4*, and *TP53* genes (2). While BTKi are effective for patients who are unsuitable for chemoimmunotherapy or have *TP53*-mutated WM, treatment is indefinite and has cumulative toxicities. The combination of chemoimmunotherapy with BTK inhibitors (BTKi) has not been widely studied (3).

“BRAWM” is a multicenter phase 2 trial looking at the combination of Bendamustine (nitrogen mustard alkylating chemotherapy) and Rituximab (anti-CD20 monoclonal antibody) with Acalabrutinib (a second-generation BTKi), termed “BR-A”, in treatment-naïve WM (clinicaltrials.gov, NCT04624906). For many WM patients, progression free survival (PFS) after initial treatment can be up to 78-82 months (4). Therefore, anticipatory evaluations of efficacy, such as minimal residual disease (MRD) analysis, are required as an adjunct to traditional clinical trial endpoints (5).

Multiple methods of MRD monitoring have been used as measures of response in WM and indolent lymphoma, including digital droplet or quantitative PCR of *MYD88* p.L265P, VDJ sequencing of the clonal B cell with clonoSEQ and targeted-capture panel sequencing (6), Techniques can be utilized in bone marrow (BM) cells or peripheral blood (PB). These approaches are limited in sensitivity due to targeting relatively few genomic alterations found in the tumor. Whole genome sequencing (WGS) of cell-free DNA (cfDNA)is a non-invasive method of tumor detection that has the advantage of detecting multiple genomic alterations and may improve MRD sensitivity by sampling thousands of mutations from across the genome rather than relying on detection of a specific alteration (7).

We assessed the utility of blood cfDNA whole genome sequencing (cfWGS) for MRD monitoring in WM through comparison to targeted panel sequencing of 1) immunoglobulin (IG) VDJ genes (CapIG-seq) 2) mutations in *MYD88* and *CXCR4* genes, and 3) clonoSEQ. All assays were tested on PB cells (clonoSEQ), cfDNA (targeted panel), and BM samples collected every 6 months for 18 months from 7 patients with WM treated uniformly on the BRAWM study.

## Materials and Methods

### Study Design

Peripheral blood (PB) and bone marrow (BM) samples were collected from consenting patients enrolled in the BRAWM study. Study participants were symptomatic, treatment-naive patients over age 18 years, with biopsy-proven WM who consented to participate in biobanking. Samples were collected every 6 months at 9 participating Canadian sites: 1) “pre-treatment”: immediately before BR-A treatment; 2) “cycle 7”: after six 4-week cycles of BR-A ; 3) “cycle 12”: after the completion of acalabrutinib-alone treatment and 4) at “18 month” follow up.

All PB and BM samples were analyzed by a targeted sequencing panel of 27 genes of interest in WM and B cell immunoglobulin gene rearrangements (Suppl.1, Suppl.2). Clinical PB and BM samples were analyzed by clonoSEQ Assay from Adaptive Biotechnologies (Seattle, WA, USA) at all timepoints. Pre-treatment BM were analyzed by a targeted clinical panel of *MYD88, CXCR4* and *TP53* at Health Sciences North (Sudbury, ON, Canada).

The BRAWM clinical trial was approved by the Ontario Cancer Research Ethics Board (OCREB, CTO project #3242). The biomarker substudy was approved by the Sunnybrook Research Ethics Board (REB#5500).

### Sample Collection and Processing

At each timepoint, 2-3mL of BM was collected in EDTA tubes along with 20-30mL of PB in Streck Cell-Free DNA BCT tubes. Samples were shipped overnight and separated into components upon receipt (within 24-28 hours of collection, exceptions noted in Table 1 and Suppl1). BM samples were separated by Ficoll density gradient centrifugation to isolate mononuclear cells (BMMC) and then selected for CD19 expression using the EasySep™ Human CD19 Positive Selection Kit as per the manufacturer’s protocol. To isolate PB plasma, Streck tubes were centrifuged at 1,600g for 10 minutes at 4C. The plasma layer was pipetted into a fresh microcentrifuge tube and centrifuged at 16,000g for 10 minutes at 4C. PBMC were retained as a matched normal control for cfWGS. Samples stored at -80C were batched for extraction and processing.

**Table 1.**
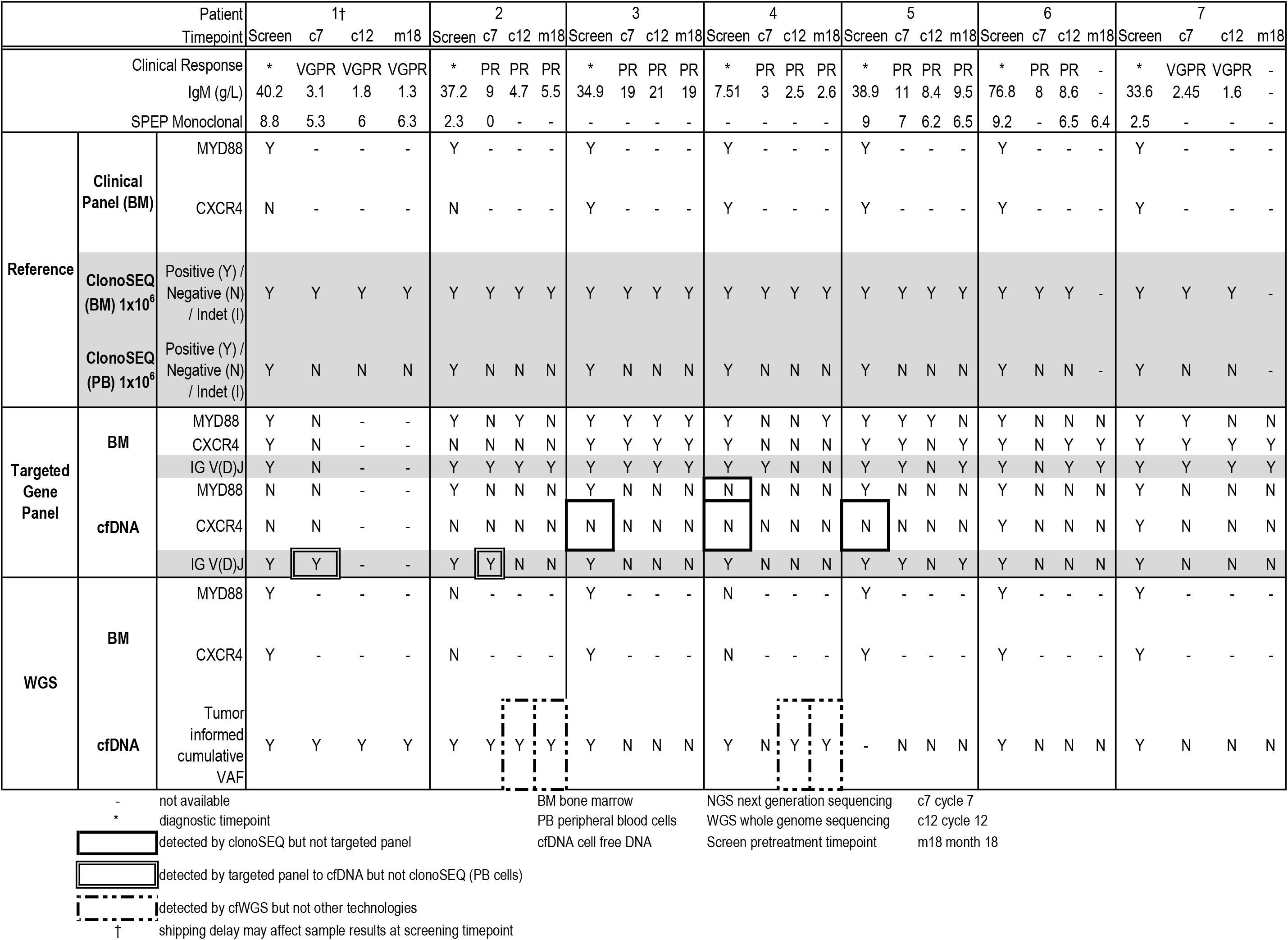

### Clinical ClonoSEQ and MYD88 Assays

1mL of BM and 10mL of PB collected in EDTA tubes at local study sites were frozen at -80C and batched for shipping to Adaptive Biotechnologies biannually (Seattle, WA, USA). Samples were thawed and processed according to a validated commercial protocol from Adaptive Biotechnologies. In brief, this protocol involved barcoded, multiplex PCR amplification of multiple B-cell immunoglobulin loci (IGH VDJ and DJ, IGK and IGL), followed by deep Illumina sequencing to identify and quantify rearranged B-cell receptor gene sequences.

1mL of BM was collected in an EDTA tube at the time of diagnosis and shipped at ambient temperature within 24 hours to Health Sciences North (HSN) genomics centre (Sudbury, ON, Canada) within 24 hours of collection. CD19+ cells were selected for DNA extraction via immune-bead technology, followed by hybrid capture and targeted sequencing of *MYD88, CXCR4* and *TP53*.

### DNA extraction and Library Construction

Following the manufacturer’s instructions, cfDNA was extracted from PB plasma using the QIAamp Circulating Nucleic Acid Kit while genomic DNA was extracted from BM and PB BC using either the Qiagen AllPrep DNA/RNA/miRNA kit or Gentra Puregene Blood Kit. Genomic DNA was sheared by ultrasonication (Covaris LE220) to 250bp median fragment length prior to library preparation. cfDNA did not require shearing. Barcoded DNA libraries were prepared using the KAPA HyperPrep kit. For WGS, these pre-capture libraries were used for Illumina sequencing directly.

### Targeted Panel Design and Capture

To enable deep coverage of specific genes, we designed a custom gene panel of 1,223 probes targeting the exons of 27 genes of interest in WM (Integrated DNA Technologies, Suppl Table 1, Suppl Table 2). These probes were combined with a set of 771 probes targeting the immunoglobulin V, D, and J genes to detect B-cell VDJ rearrangements, termed “CapIG-seq” (8). Target capture was performed as previously described, following the Integrated DNA Technologies, xGEN protocol (8).

### Sequencing

Pre-capture WGS and post-capture targeted panel libraries were sequenced using the Illumina NovaSeq X Plus using paired-end 150 bp reads. Targeted panel libraries were sequenced to a target median depth of 3000-5000X regardless of sample type. WGS libraries were sequenced to a target depth of 80X for CD19+ selected BM, 30X for PB BC normal samples and 40X for cfDNA.

### Bioinformatic Analysis

#### Targeted Panel Analysis

Targeted sequencing reads were aligned to the human reference genome (hg38) using the Burrows-Wheel Aligner (BWA, v0.7.15) available from https://github.com/lh3/bwa/releases. Indelrealigner(gatk/4.1.8.1)(https://gatk.broadinstitute.org/hc/en-us/sections/360009803432-4-1-8-1) was run as post-processing of the bam files. Error correction was achieved using molecular adaptors to form consensus sequences using the ConsensusCruncher (9). Candidate mutations were called using muTect2 (gatk/4.1.8.1) (10), followed by filtration of germline variants present in the population from the gnomAD v2.1 database(11) and in-house panel of 263 normals. Annotation was performed by Ensembl Variant Effect Predictor(v92) (12). We used the MANE Select transcripts *CXCR4* (NM_003467.3), *TP53*(NM_000546.5) and *MYD88* (NM_002468.5) for mutation calling and recoded the MYD88 p.L252P to p.L265P to be consistent with the literature. To focus on high-quality variant calls, we removed the mutations that were rejected by muTect2, plus all point variants <1% variant allele frequency (VAF) and in/dels <5% VAF. For MRD analysis below these thresholds, we manually verified that *MYD88* p.L265P and *CXCR4* variant calls matched the exact mutations present in the baseline sample.

MiXCR (3.0.13) (13) was used for V(D)J immunoglobulin rearrangement calling. Outputs were verified to contain at least one productive rearrangement in *IGH, IGK* or *IGL*. Candidate rearrangements were filtered for a minimum clonal fraction of 5% and a clone count of 50 in diagnostic BM. In follow-up samples, the presence of the diagnostic clone at any level was sufficient for samples to be considered MRD positive. (Suppl. Table 3)

#### Whole Genome Sequencing Analysis

WGS reads were aligned to the human reference genome (hg38) using the Ontario Institute for Cancer Research (OICR) *bwaMem* pipeline (v1.0.0; [GitHub](https://github.com/oicr-gsi/bwaMem) https://github.com/oicr-gsi), incorporating Burrows-Wheel Aligner (BWA, v0.7.12), samtools (v1.9), cutadapt (v1.8.3), and other dependencies under default parameters. Bam files were further processed through coordinate sorting (*samtools* v1.9), duplicate marking (*Picard* v2.19.2), local realignment around indels (*GATK* IndelRealigner), and base quality score recalibration (*GATK* BaseRecalibrator), detailed in the OICR *bamMergePreprocessing* workflow (v2.1.1; [GitHub](https://github.com/oicr-gsi/bamMergePreprocessing)). Somatic single-nucleotide variants (SNVs) and small insertions/deletions (indels) were called from paired tumour/normal WGS data using the OICR *mutect2* pipeline (v1.0.9; [GitHub](https://github.com/oicr-gsi/mutect2)).

Next, we filtered somatic mutations from baseline BM WGS to define high-confidence somatic variants for MRD tracking. Mutations were retained if they had a VAF ≥10%, passed GATK’s FilterMutectCalls, did not overlap ENCODE Blacklist(15) or RepeatMasker regions(16), and did not overlap known germline variants (any variant with a dbSNP identifier). These patient-specific mutation lists were used as input to a modified version of MRDetect (7), which calculates a detection rate based on the proportion of reads supporting known baseline mutations relative to total reads at those sites. A z-score was then calculated based on the number of mutant sites detected in cfDNA relative to a healthy donor cohort (n = 25), with a z-score ≥4.5 classified as MRD-positive.

## Results

Clinically, all patients in this cohort achieved and sustained a very good partial response (VGPR) or partial response (PR) by the Sixth International Workshop on Waldenstrom’s Macroglobulinemia (IWWM) criteria (Table 1) (1).

### MYD88 and CXCR4 mutation detection

We first compared mutation detection between the clinical panel and our targeted sequencing panel on BM and cfDNA samples at the timepoint of highest mutation burden, pre-treatment. Clinical panel analysis of pre-treatment BM established that all 7 patients carried cancers with *MYD88* p.L265P mutation and 5/7 also harboured *CXCR4* mutations (Table 1). We verified all of these mutations by targeted panel sequencing as well as one additional *CXCR4* p.S341Ffs*3 mutation in the C-terminal domain not detected by clinical testing (Table 1, Patient 1). However, the same panel applied to pre-treatment cfDNA only detected 5/7 *MYD88* mutations and found only 2/6 *CXCR4* mutations (Table 1, Suppl2). These data suggest poorer sensitivity of the targeted gene panel in cfDNA compared to BM.

Next, we compared MRD in BM measured by clonoSEQ with the results of targeted panel sequencing of *MYD88* or *CXCR4* in BM or cfDNA. With clonoSEQ BM, all patients remained positive throughout treatment. Using targeted panel sequencing, 1 patient remained positive for *MYD88* p.L265P throughout, and 6 patients became MRD negative: 4 by cycle 7, 1 by cycle 12 and 1 at month 18. We observed the reemergence of MRD in 2 patients: 1 at cycle 12 and 1 at month 18. In cfDNA, all 5 patients became MRD negative by cycle 7 and remained negative. Overall, both blood-based techniques were inadequate to detect persistent disease in BM.

### Immunoglobulin detection

Before treatment, all patients were positive for immunoglobulin VDJ by clonoSEQ in PB cells and BM. All patients remained clonoSEQ positive in BM throughout treatment; however, in PB cells, all patients became clonoSEQ negative at 7 months and remained negative. By CapIG-seq of BM, 3/7 patients were positive throughout their treatment course, 1/7 patients became negative by cycle 7, 1/7 became negative by cycle 12 and stayed negative, and 2/7 patients became negative and then had reemergence of MRD at 18 months (Table 1, patients 5 and 6). By comparison, CapIG-seq of cfDNA found that 4/7 patients became negative at cycle 7, the remaining 3/7 patients also tested negative by 12 months, and no patients had a reemergence of MRD. Overall, cfDNA CapIG-seq was more sensitive than PB cell clonoSEQ for MRD detection (ρ = 0.78, Figure 2).

### Tumour-informed cfWGS and comparison with orthogonal MRD technologies

We performed tumour-informed PB cfWGS on 6 patients at all timepoints and 1 patient from cycle 7 and beyond due to a lack of residual cfDNA. Somatic mutation lists were generated from CD19-selected bone marrow samples (mean coverage 57X; SD 24; range 45–68) and matched normal PBMCs (mean coverage 21X; SD 4; range 21–29) after standard preprocessing (see Methods). On average, we called 1,300 somatic mutations per cancer genome with a standard deviation of 564 (range 355–1813). These mutations included the 5 MYD88 somatic mutations at >0.5% VAF detected by targeted panels, but missed the 2 mutations below that level (Suppl Table 2).

To sample these mutations over time as a measure of MRD, we sequenced the PB cfDNA samples to a mean target depth of 31X (SD 8, range 13-45) WGS. In the 6 samples taken pre-treatment, we detected a statistically significant number of mutations, 42-845 per patient (median = 151), which corresponded to an aggregate tumour fraction of 0.6%-7.4% (median = 1.9%) (Figure 1A). These tumour fractions measured by cfWGS correlated strongly with those measured by targeted panel for *MYD88* in bone marrow (Spearman’s ρ = 0.93, p = 0.008), but showed only weak, non-significant associations with *MYD88* VAFs in cfDNA (ρ = 0.31, p = 0.564), clonoSEQ in bone marrow (ρ = 0.09, p = 0.919), and clonoSEQ in peripheral blood (ρ = –0.43, p = 0.419) (Figure 2). In the cycle 7 samples, we observed a sharp decline in tumour fraction with a statistically significant number of mutations detected in only 2/7 patients (91 and 13) and at low aggregate tumour fractions (0.59% and 0.85%). We classified the remaining 5 patients as MRD negative, consistent with clonoSEQ in cfDNA but not clonoSEQ in BM, where all 5 patients were positive (Figure 1A, 2).

**Figure 1.**
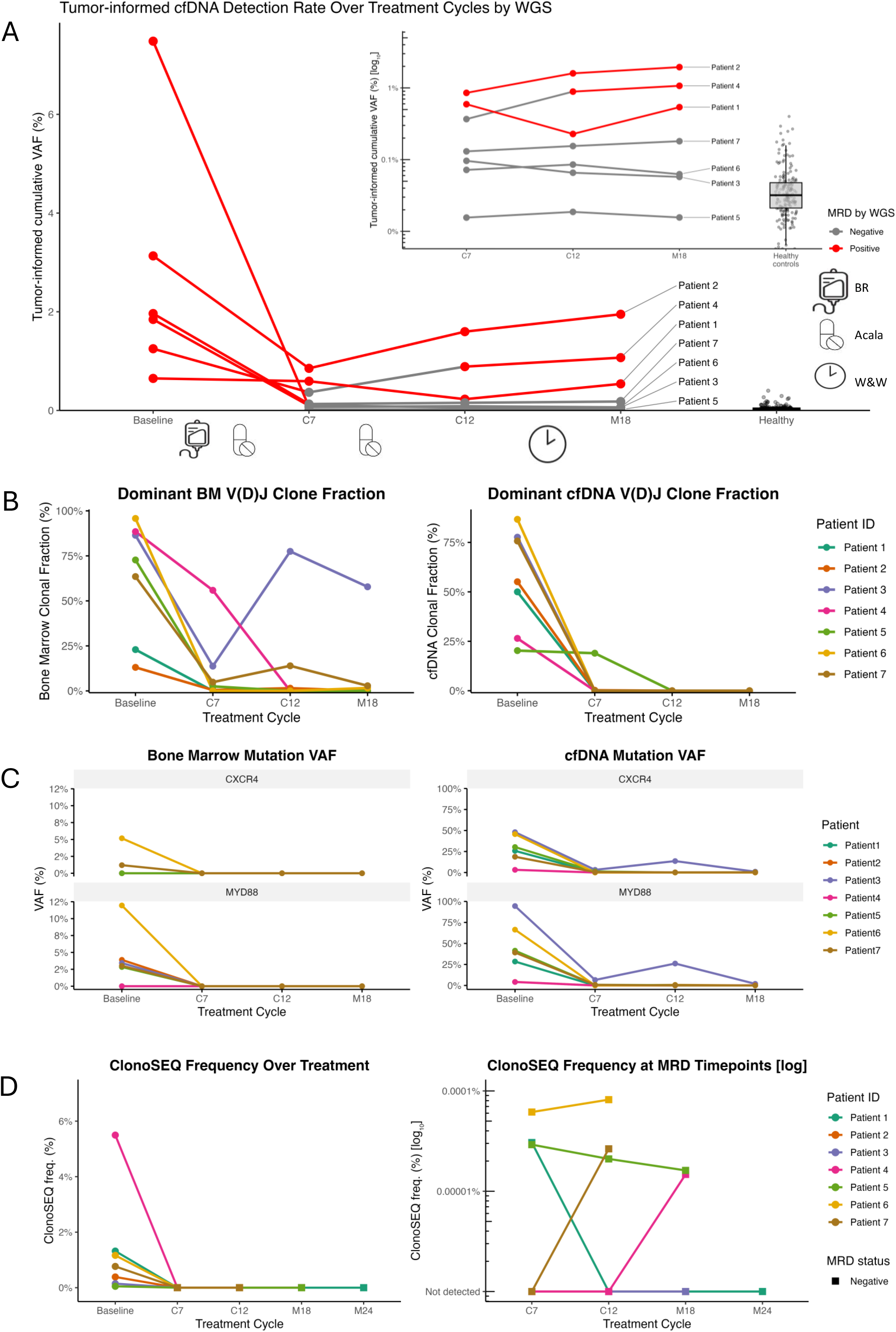
Longitudinal profiling of tumor burden and clonal dynamics in patients with Waldenström’s macroglobulinemia. **(A)** Tumor-informed cfDNA detection by whole-genome sequencing (WGS), shown as cumulative variant allele fraction (VAF %) over time. Lines represent individual patients at baseline (post–bendamustine–rituximab–acalabrutinib), after cycle 7 of acalabrutinib (C7), and at follow-up timepoints (Cycle 12 and Month 18). MRD status by WGS is shown (red = positive, gray = negative); healthy donor controls are plotted at right. The insert shows the same data on a log_10_ scale. Samples with a significantly higher proportion of tumor-somatic variants relative to healthy controls are marked as MRD-postivie. (B) Dominant V(D)J clonal fractions in bone marrow (left) and cfDNA (right) for each patient, tracking the highest Baseline clone across timepoints. (C) Allele fractions for CXCR4 and MYD88 mutations in bone marrow (left) and cfDNA (right), assessed by targeted NGS. (D) ClonoSEQ MRD frequencies over time, shown on linear (left) and log10 (right) scales for the treatment timepoints. Shapes denote MRD status (circle = positive, square = negative, triangle = indeterminate).

**Figure 2.**
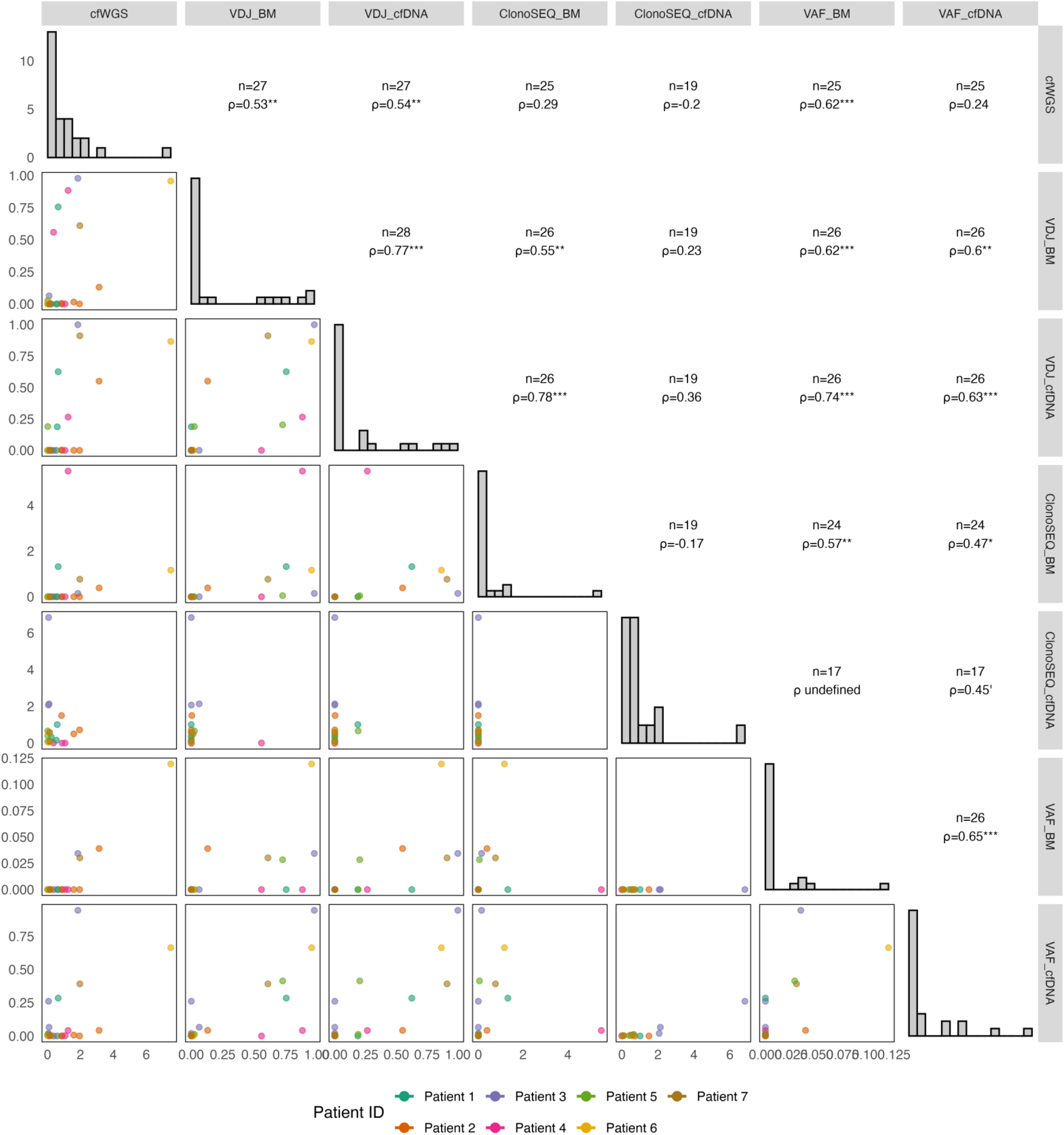
Pairwise relationships among multimodal tumor-burden metrics. Lower panels show scatterplots of six longitudinal metrics for each patient (n indicated in upper panels): cfWGS_detection_rate (cumulative cfDNA detection rate by WGS as a percent), VDJ_BoneMarrow (the clonal fraction of the dominant V(D)J clone known from baseline across longitudinal BM samples) and VDJ_cfDNA (the proportion of the dominant V(D)J clonal fraction in cfDNA), ClonoSEQ_pct (ClonoSEQ-derived tumor frequency %, plotted as one divided by the per million cell count), BM_cells_target_VAF and cfDNA_target_VAF (maximal targeted-NGS mutation VAF in CXCR4/MYD88 in BM cells or cfDNA). Points are colored by patient (1–7). Diagonal panels display the overall distribution of each metric. Upper panels report Spearman’s ρ (with significance: *p < 0.05; **p < 0.01; ***p < 0.001) and two-sided p-values for each pairwise comparison. This matrix highlights the strength of concordance between orthogonal assays of minimal residual disease across treatment timepoints.

In subsequent timepoints, the 2 MRD-positive patients remained positive and displayed increasing aggregate tumour fractions (0.23% to 0.54% for Patient 1 and 1.60% to 1.95% for Patient 2 from cycle 12 to month 18). Of the 5 patients MRD negative at cycle 7, one patient subsequently became MRD positive with significant numbers of mutations detected (30/613 at cycle 12, compared to 15 detected at cycle 7) and increasing tumour fractions (0.086% to 0.18% from cycle 7 to cycle 12) (Figure 1A). Compared to the targeted panel for *MYD88* and CapIg-seq, cfWGS was positive for two patients at both cycle 12 and month 18, where other targeted PB technologies failed to detect MRD (Table 1, Figure 1 A-C). These positives were all confirmed by clonoSEQ in BM, validating cfWGS results (Table 1). While all PB technologies demonstrated a drop in mutation/IG burden at cycle 7, cfWGS was most sensitive in detecting the re-emergence of MRD (Figure 1).

## Discussion

In this pilot cohort of seven WM patients treated on a novel chemoimmunotherapy + BTKi regimen, we compared MRD detection technologies applied to BM and PB cells and cfDNA. In addition to well-established targeted panel, CapIG-seq, and clonoSEQ protocols, this comparison included a new, tumour-informed cfWGS mutational sampling (40X target coverage) approach pioneered in high mutation rate tumours but not previously applied to lower mutation rate WM (7,17).

We observed poor sensitivity for both clonoSEQ (PB) and our custom targeted panel (cfDNA) to detect residual WM, compared to persistent positivity in the corresponding BM for both techniques. In these seven patients, cfWGS appeared to outperform both techniques when compared to BM clonoSEQ. These preliminary results require validation in larger cohorts. PB cell MRD has previously been demonstrated to be an independent predictor of outcomes in WM, as MRD-positive patients have comparatively poorer outcomes (18,19). Molecular markers have demonstrated superiority to traditional IgM measures due to the significant component of WM B-cells that may not produce immunoglobulin (20). Use of cfWGS as a more sensitive marker of MRD may be useful to further improve prognostic models for WM. Long-term follow-up and evaluation in a larger sample size are needed to validate this.

Most patients with WM enjoy long periods of stability between treatments, where clinical management consists of routine surveillance, “watch and wait”. However, the International WM Working group (IWWM) recognizes that there are subsets of patients that behave more aggressively, “high risk smouldering WM” or “high risk WM” (HRSWM/HRWM) (21) and recommends close clinical surveillance. Currently, these subgroups are defined by clinical risk stratification systems, though molecular markers are becoming more clearly defined (22). Mutations of *CXCR4, MYD88* and *TP53* are the most well-established biomarkers for predicting a more aggressive course (2), however, this study demonstrates the difficulty with routine PB monitoring as single gene testing in PB may correlate poorly to BM when the disease burden is low. As routine BM is not typically recommended during expectant management, cfWGS may represent a promising non-invasive tool to evaluate the need for further investigation in this high-risk group. This proof-of-principle also suggests that an appreciable fraction of the WM genome is accessible via cfDNA and that deeper profiling may enable in-depth mutation calling and detection of relapse mechanisms in longitudinal samples.

## Data Availability

Primary whole-genome sequencing data are not available as the patients did not provide explicit written permission for their complete sequence to be shared. Secondary variant calls are available in the supplementary tables, as are the clinical data used for figures. The source code needed to reproduce the figures is available at https://github.com/pughlab/wm-cfwgs-mrd/.

## Author Contributions

The study was designed by S. Chow, N. Berinstein, C. Chen, S. Trudel and T.J. Pugh. S. Chow drafted the manuscript with significant input from T.J. Pugh and D. Abelman. D. Abelman conducted bioinformatics analyses, generated manuscript visualizations, and performed the statistical analyses. A. Danesh conducted bioinformatic analysis. K. Roos provided logistical support for clinical trial samples and coordination with clinical partners. E. N. Wei and S. Pedersen conducted the laboratory work, including DNA extractions, library preparation and targeted capture. A. Suleman provided support with analysis from the clinical assays. D. Scott provided critical manuscript support. All authors read, provided input and approved the manuscript.

## Acknowledgements

Funding for this research is provided by the Robert A. Kyle Career Development Award from the International Waldenstrom’s Macroglobulinemia Foundation (IWMF), Waldenstrom’s Macroglobulinemia Foundation of Canada and Poh Family Research Fund to S. Chow.

D. Ableman is supported by the Princess Margaret Cancer Foundation Graduate Fellowship in Cancer Research, the Dr. J.R. Cunningham Graduate Fellowship in Cancer Research, and the Princess Margaret Hospital Foundation Graduate Fellowship in Cancer Research.

T.J. Pugh holds the Canada Research Chair in Translational Genomics and is supported by a Senior Investigator Award from the Ontario Institute for Cancer Research and the Gattuso-Slaight Personalized Cancer Medicine Fund.

The authors are grateful for the technical support of the Ontario Institute for Cancer Research Genomics Program (https://genomics.oicr.on.ca, supported by the Government of Ontario), the Princess Margaret Genomics Center (www.pmgenomics.ca), UHN HPC and Bioinformatics core, and Centre for Clinical Trial Support (CCTS) at Sunnybrook Research Institute (SRI) (https://research.sunnybrook.ca/research/centre-for-clinical-trial-support-ccts/).

Adaptive Biotechnologies provided in-kind support for clonoSEQ assays. The BRAWM study is sponsored by AstraZeneca.

